# Disparities in Mortality Associated with Acute Myocardial Infarction and COVID-19 in the United States: A Nationwide Analysis

**DOI:** 10.1101/2023.03.30.23287987

**Authors:** Amer Muhyieddeen, Susan Cheng, Mamas A Mamas, Dorian Beasley, Galen Cook Weins, Martha Gulati

**Author notes:** CORRESPONDENCE: Martha Gulati, MD, MS, FACC, FAHA, FASPC, FESC, 127 S. San Vicente Blvd, Suite A3600, Los Angeles, CA 90048. Disclosures: M.G. served on an advisory board for Novartis and Esperion. She is a co-investigator and site PI of the Women’s IschemiA TRial to Reduce Events In Non-ObstRuctive CAD (WARRIOR) Study funded by the Department of Defense (Award Number: W81XWH- 17-2-0030).

## Abstract

**Background:** The impact of the COVID-19 pandemic on potential racial disparities in acute myocardial infarction (AMI) management and outcomes is unclear. We examined AMI patient management and outcomes during the pandemic’s initial nine months, comparing COVID-19 and non-COVID-19 cases.

**Methods:** We identified all patients hospitalized for AMI in 2020 using the National Inpatient Sample (NIS), identifying those with or without concurrent COVID-19. Logistic and linear regression was used for analyses of associations, with adjustment for potential confounders.

**Results:** Patients with both AMI and COVID-19 had higher in-hospital mortality rates (aOR 3.19, 95% CI 2.63-3.88), mechanical ventilation (aOR 1.90, 95% CI 1.54-2.33), and hemodialysis (aOR 1.38, 95% CI 1.05-1.89) compared to those without COVID-19. Black and Asian/Pacific Islander patients had higher in-hospital mortality than White patients, (aOR 2.13, 95% CI 1.35-3.59) and (aOR 3.41, 95% CI 1.5-8.37). Moreover, Black, Hispanic, and Asian/Pacific Islander patients had higher odds of initiating hemodialysis, (aOR 5.48, 95% CI 2.13-14.1), (aOR 2.99, 95% CI 1.13-7.97), and (aOR 7.84, 95% CI 1.55-39.5) and were less likely to receive PCI for AMI, (aOR 0.71, 95% CI 0.67-0.74), (aOR 0.81, 95% CI 0.77-0.86), and (aOR 0.82, 95% CI 0.75-0.90). Additionally, Black patients had a lower likelihood of undergoing CABG surgery for AMI (aOR 0.55, 95% CI 0.49-0.61).

**Conclusion:** Our study revealed increased mortality and complications in COVID-19 patients with AMI, highlighting significant racial disparities. Urgent measures addressing healthcare disparities, such as enhancing access and promoting culturally sensitive care, are needed to improve health equity.

## Introduction

The COVID-19 pandemic significantly strained healthcare systems worldwide once it was declared a pandemic by the World Health Organization in March 2020. As of January 5, 2023, over 638 million people had been infected, resulting in more than 6.6 million deaths.^1^ Notably, an increase in mortality rates related to cardiovascular disease has been observed during the pandemic.^2, 3^ The underlying causes of this trend remain uncertain, but potential factors include delayed or deferred care and varying treatment approaches due to hospital capacity constraints.^4^

Several studies comparing acute myocardial infarction (AMI) management and outcomes before and during the pandemic revealed longer symptom-to-balloon times, decreased adherence to medically-guided therapy, and increased mortality rates.^5–8^ Recent data also underscores the guarded outcomes of patients hospitalized for AMI and COVID-19.^9–11^ In a study analyzing clinical, procedural, and in-hospital prognostic factors for patients with COVID-19 admitted with a diagnosis of ST-elevation myocardial infarction (STEMI), higher rates of stent thrombosis, cardiogenic shock, and in-hospital mortality were reported in patients infected with COVID-19 compared to non-COVID-19 STEMI patients.^12^

Beyond the direct effects of COVID-19 on patients with cardiovascular disease, the pandemic has led healthcare and research organizations to shift resources away from non-COVID-related issues, severely impacting patients with cardiovascular conditions.^13^ This has impacted certain groups in society more than others, including socio-economically disadvantaged patient groups, and Black and other minority racial groups.^14–16^ Current literature is limited in exploring potential disparities in AMI treatment or heightened mortality among minority racial groups during the pandemic. To address this knowledge gap, our study utilized the National Inpatient Sample (NIS) to compare clinical outcomes in patients diagnosed with AMI, both with and without COVID-19 infection, and to investigate potential racial disparities in treatment and outcomes.

## Methods

The analysis was conducted using the NIS database for the year 2020. The NIS is part of the healthcare cost and utilization project (HCUP) databases and is sponsored by the agency for healthcare research and quality (AHRQ). It contains clinical and resource utilization information on millions of discharges annually, with precautions to safeguard the privacy of individual patients and hospitals. The data is stratified to represent 20% of U.S. inpatient hospitalizations across different hospitals and geographic areas as a random sample. The NIS database allows for calculating national estimates by providing a weight variable.^17^ For 2020, the unweighted sample included 6.3 million observations, and the weighted sample was around 31.7 million discharges. All patients admitted to the hospital with acute myocardial infarction [STEMI or non-ST-elevation myocardial infarction (NSTEMI)], and concomitant COVID-19 infection were included in this study. Because our study used deidentified data, it was exempt from Institutional Review Board approval.

To identify patients admitted with AMI, the NIS database was searched using the International Classification of Diseases, Tenth Revision, Clinical Modification (ICD-10-CM) codes (I20.0, I21.1, I21.2, I21.3, and I21.4). In the present analysis, a total of 16,465 cases were excluded on account of elective admissions. Furthermore, 85,174 cases were removed from the dataset to avoid duplicate counting, as these patients were transferred out of the hospital. Exclusions were also made for cases with missing variables, including insurance status, race, sex, death status, and age. The missing cases constituted less than 0.8% (3,528/446,834) of the initial dataset (Figure 1).

**Figure 1:**
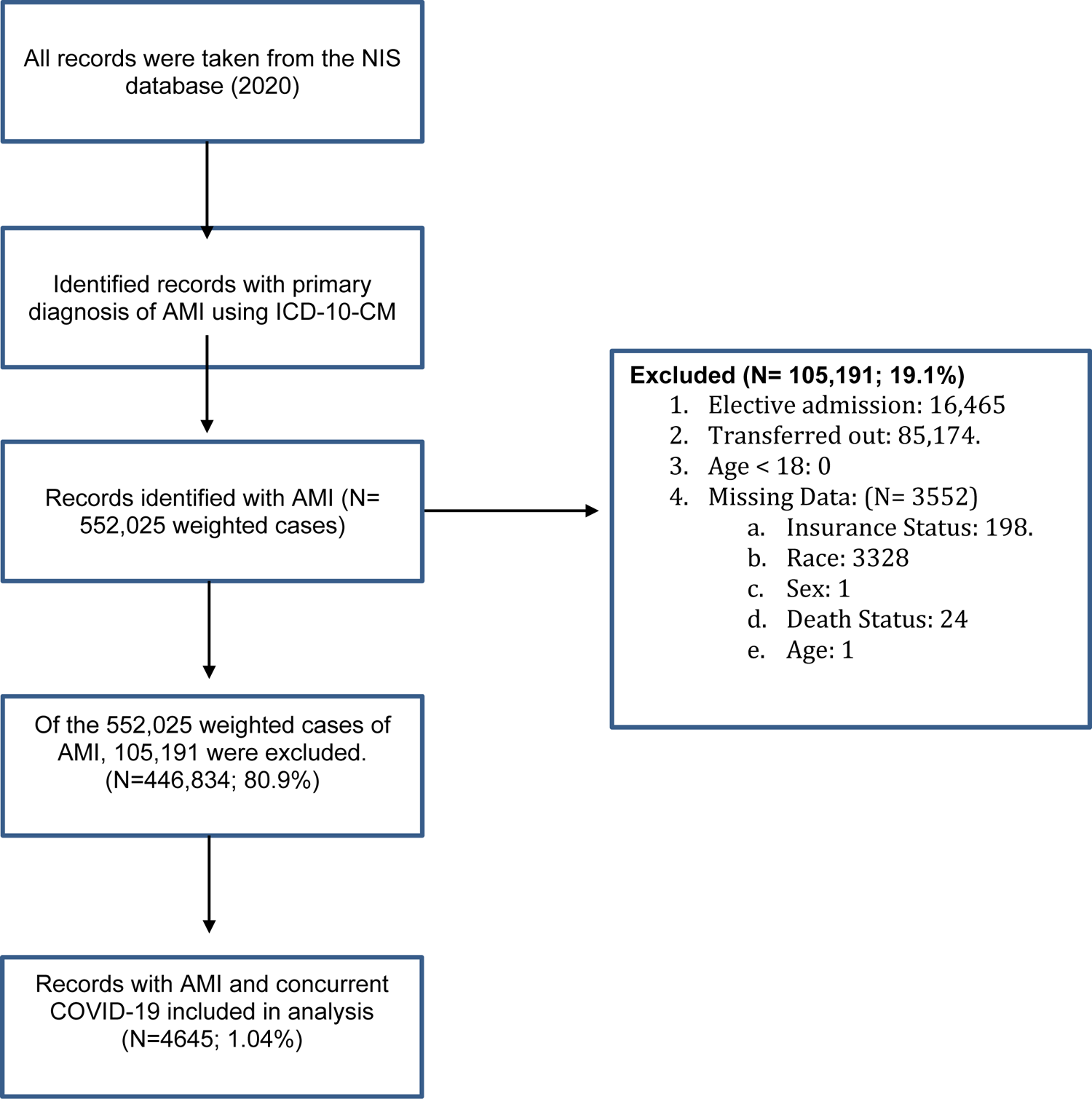
Consort Flow Diagram Depicting Inclusion and Exclusion Criteria for Study

The cohort of patients with AMI was stratified into two subgroups based on the presence or absence of COVID-19, as identified by the ICD-10 code (U07.1). The primary endpoint of the study was in-hospital mortality. Secondary outcomes encompassed revascularization rates using percutaneous coronary intervention (PCI), coronary artery bypass grafting (CABG), and thrombolytic therapy, acute kidney injury necessitating hemodialysis, administration of in-hospital vasopressors, utilization of mechanical ventilation and mechanical circulatory support, as well as the duration of hospital stay.

Statistical analysis was conducted using STATA 17 (StataCorp LLC, College Station, TX). Categorical variables were compared using chi-square tests, while linear regression was employed for continuous variables. To assess associations and adjust for potential confounders, logistic and linear regression were utilized. Candidate variables underwent testing for univariable associations, with those exhibiting a p-value <0.2 being incorporated into the final multivariable model. The Elixhauser Comorbidity Index was employed to account for comorbid conditions.^18^ We defined significance as a 2-tailed *P* value of 0.05.

## Results

During 2020, there were a total of 446,834 hospitalizations for AMI, and 4,645 were COVID-positive (1.04%). Patients with AMI and COVID-19 were younger than those without COVID- 19 (63.9 years vs. 65.4 years, p<0.01). Patients with AMI and COVID-19 were more likely to be from minority groups, with a greater proportion of Black (17.2% vs. 9.2%), Hispanic (22.4% vs. 8.9%), and Native American patients (1.7% vs. 0.51%); (p<0.01). They were also more likely to have a household income of less than $50,000 (38.1% vs. 30.7%, p= <0.01), and to have Medicaid insurance (13.4% vs. 10.5%, p< 0.01).

Patients with AMI and COVID-19 had a higher prevalence of type II diabetes (41.7% vs. 31.7%, p < 0.01), and ischemic stroke (2.3% vs. 0.9%, p < 0.01) but were less likely to have a history of coronary artery disease (81.2% vs. 86.2%, p= < 0.01), hyperlipidemia (59.9% vs. 68.1%, p<0.01), and tobacco use (35.9% vs. 53.4%, p< 0.001), compared to those with AMI without COVID-19 (Table 1).

**Table 1:**
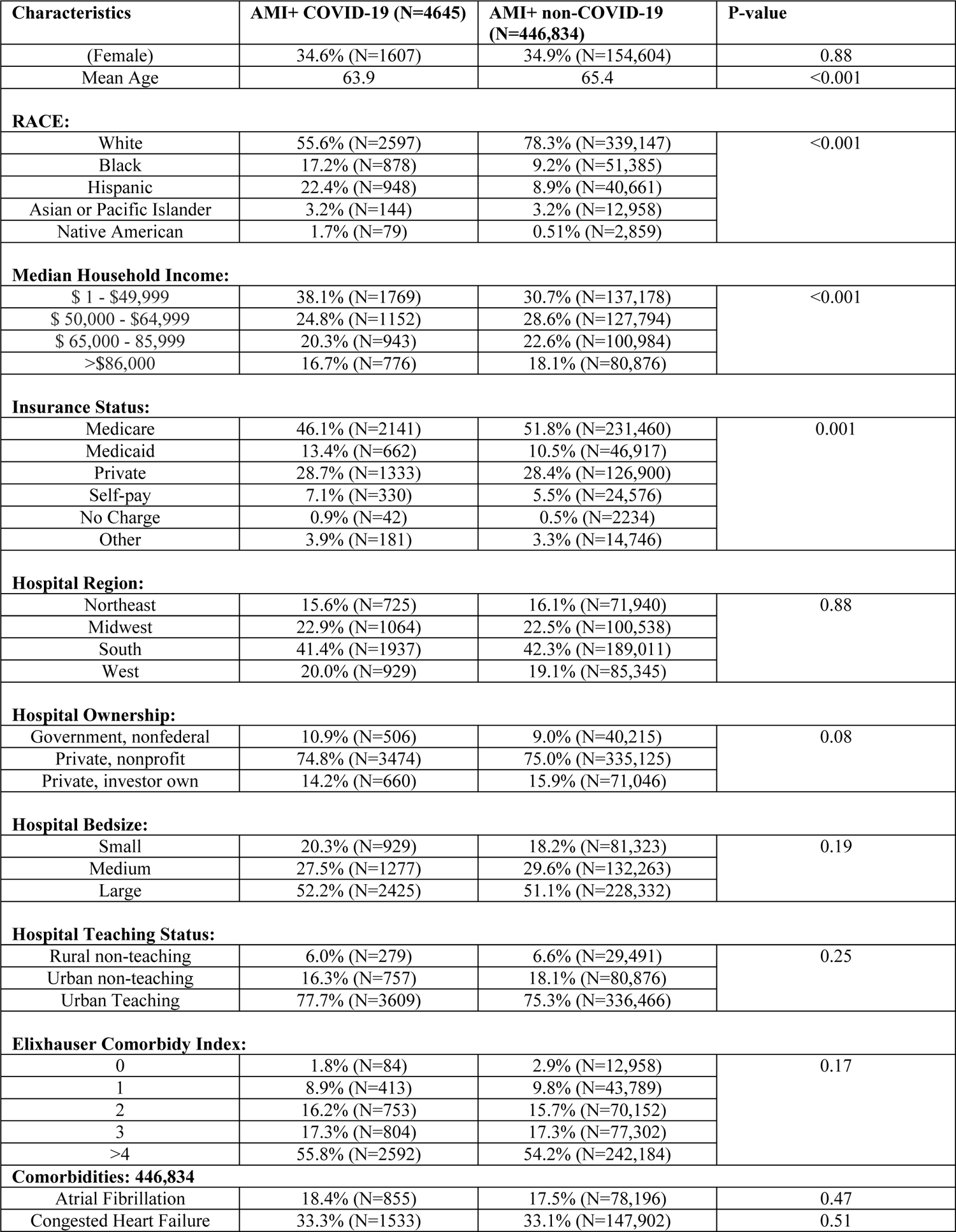
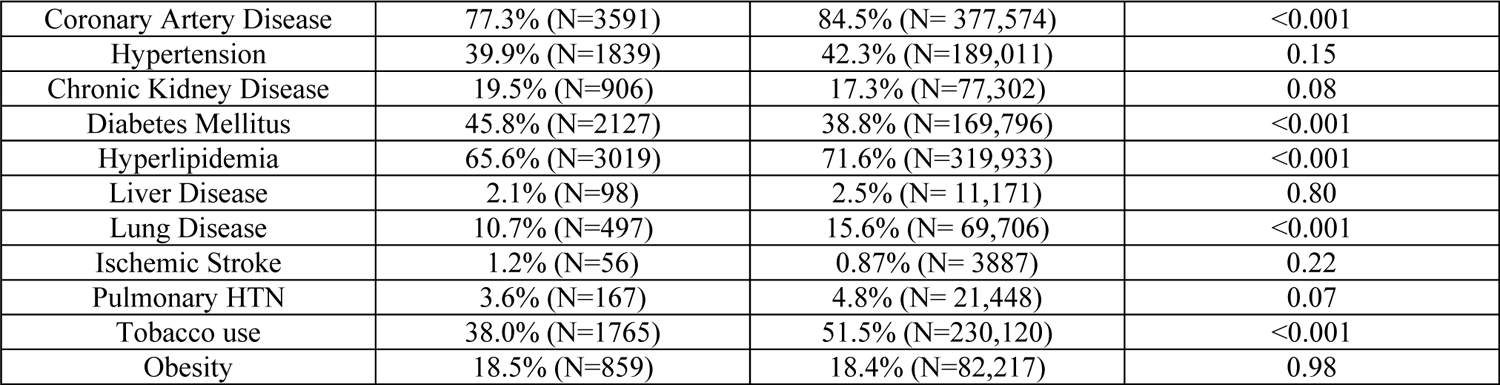
Baseline characteristics of All Acute Myocardial Infarctions based on presence or absence of COVID-19.

| Characteristics | AMI+ COVID-19 (N=4645) | AMI+ non-COVID-19 (N=446,834) | P-value |
| --- | --- | --- | --- |
| (Female) | 34.6% (N=1607) | 34.9% (N=154,604) | 0.88 |
| Mean Age | 63.9 | 65.4 | <0.001 |
| RACE: |  |  |  |
| White | 55.6% (N=2597) | 78.3% (N=339,147) | <0.001 |
| Black | 17.2% (N=878) | 9.2% (N=51,385) |  |
| Hispanic | 22.4% (N=948) | 8.9% (N=40,661) |  |
| Asian or Pacific Islander | 3.2% (N=144) | 3.2% (N=12,958) |  |
| Native American | 1.7% (N=79) | 0.51% (N=2,859) |  |
| Median Household Income: |  |  |  |
| \$ 1 - \$49,999 | 38.1% (N=1769) | 30.7% (N=137,178) | <0.001 |
| \$ 50,000 - \$64,999 | 24.8% (N=1152) | 28.6% (N=127,794) | |
| \$ 65,000 - 85,999 | 20.3% (N=943) | 22.6% (N=100,984) | |
| >\$86,000 | 16.7% (N=776) | 18.1% (N=80,876) | |
| Insurance Status: |  |  |  |
| Medicare | 46.1% (N=2141) | 51.8% (N=231,460) | 0.001 |
| Medicaid | 13.4% (N=662) | 10.5% (N=46,917) |  |
| Private | 28.7% (N=1333) | 28.4% (N=126,900) |  |
| Self-pay | 7.1% (N=330) | 5.5% (N=24,576) |  |
| No Charge | 0.9% (N=42) | 0.5% (N=2234) |  |
| Other | 3.9% (N=181) | 3.3% (N=14,746) |  |
| Hospital Region: |  |  |  |
| Northeast | 15.6% (N=725) | 16.1% (N=71,940) | 0.88 |
| Midwest | 22.9% (N=1064) | 22.5% (N=100,538) |  |
| South | 41.4% (N=1937) | 42.3% (N=189,011) |  |
| West | 20.0% (N=929) | 19.1% (N=85,345) |  |
| Hospital Ownership: |  |  |  |
| Government, nonfederal | 10.9% (N=506) | 9.0% (N=40,215) | 0.08 |
| Private, nonprofit | 74.8% (N=3474) | 75.0% (N=335,125) |  |
| Private, investor own | 14.2% (N=660) | 15.9% (N=71,046) |  |
| Hospital Bedsize: |  |  |  |
| Small | 20.3% (N=929) | 18.2% (N=81,323) | 0.19 |
| Medium | 27.5% (N=1277) | 29.6% (N=132,263) |  |
| Large | 52.2% (N=2425) | 51.1% (N=228,332) |  |
| Hospital Teaching Status: |  |  |  |
| Rural non-teaching | 6.0% (N=279) | 6.6% (N=29,491) | 0.25 |
| Urban non-teaching | 16.3% (N=757) | 18.1% (N=80,876) |  |
| Urban Teaching | 77.7% (N=3609) | 75.3% (N=336,466) |  |
| Elixhauser Comorbidity Index: |  |  |  |
| 0 | 1.8% (N=84) | 2.9% (N=12,958) | 0.17 |
| 1 | 8.9% (N=413) | 9.8% (N=43,789) |  |
| 2 | 16.2% (N=753) | 15.7% (N=70,152) |  |
| 3 | 17.3% (N=804) | 17.3% (N=77,302) |  |
| >4 | 55.8% (N=2592) | 54.2% (N=242,184) |  |
| Comorbidities: 446,834 |  |  |  |
| Atrial Fibrillation | 18.4% (N=855) | 17.5% (N=78,196) | 0.47 |
| Congested Heart Failure | 33.3% (N=1533) | 33.1% (N=147,902) | 0.51 |
| Coronary Artery Disease | 77.3% (N=3591) | 84.5% (N= 377,574) | <0.001 |
| Hypertension | 39.9% (N=1839) | 42.3% (N=189,011) | 0.15 |
| Chronic Kidney Disease | 19.5% (N=906) | 17.3% (N=77,302) | 0.08 |
| Diabetes Mellitus | 45.8% (N=2127) | 38.8% (N=169,796) | <0.001 |
| Hyperlipidemia | 65.6% (N=3019) | 71.6% (N=319,933) | <0.001 |
| Liver Disease | 2.1% (N=98) | 2.5% (N= 11,171) | 0.80 |
| Lung Disease | 10.7% (N=497) | 15.6% (N= 69,706) | <0.001 |
| Ischemic Stroke | 1.2% (N=56) | 0.87% (N= 3887) | 0.22 |
| Pulmonary HTN | 3.6% (N=167) | 4.8% (N= 21,448) | 0.07 |
| Tobacco use | 38.0% (N=1765) | 51.5% (N=230,120) | <0.001 |
| Obesity | 18.5% (N=859) | 18.4% (N=82,217) | 0.98 |

Unadjusted clinical outcomes showed that patients with both AMI and COVID-19 had significantly higher in-hospital mortality rates (14.7% vs. 5.7%, p < 0.01), increased use of mechanical ventilation (12.8% vs. 7.4%, p < 0.01), more frequent vasopressor administration (3.6% vs. 2.2%, p < 0.01), and a greater need for hemodialysis (4.7% vs. 3.1%, p < 0.001) compared to patients with AMI alone. Additionally, their hospital stays were significantly longer, averaging 5.18 days compared to 3.83 days for those with only AMI (p<0.01). Revascularization also varied between the two groups. Patients with both AMI and COVID-19 had lower rates of PCI (39.8% vs. 45.8%, p < 0.001), combined PCI or thrombolytic usage (40.5% vs. 46.1%, p < 0.01), and CABG surgery (3.3% vs. 7.9%, p < 0.001). However, they had higher rates of thrombolytic usage (10.8% vs. 5.5%, p = 0.032) compared to patients with AMI but no COVID-19 (Table 2 and Supplemental Figure 1).

**Table 2:** Unadjusted primary and secondary outcomes of patients with AMI+ COVID-19 vs AMI without COVID-19

|  | **AMI+COVID-19** | **AMI+ non-COVID-19** | **P-value** |
| --- | --- | --- | --- |
| **In-hospital mortality** | 14.7% | 5.7% | <0.001 |
| **Mechanical ventilation** | 12.8% | 7.4% | <0.001 |
| **Vasopressor use** | 3.6% | 2.2% | <0.01 |
| **Mechanical Circulatory Support** | 1.2% | 1.3% | 0.89 |
| **Hemodialysis** | 4.7% | 3.1% | <0.01 |
| **PCI** | 39.8% | 45.8% | <0.001 |
| **Thrombolytics** | 10.8% | 5.5% | 0.032 |
| **PCI or Thrombolytics** | 40.5% | 46.1% | <0.01 |
| **CABG** | 3.3% | 7.9% | <0.001 |
| **Mean length of stay (days)** | 5.18 | 3.83 | <0.001 |
| **Total Charges (USD)** | 110,163 | 109,798 | 0.44 |

After adjusting for confounders, patients with AMI and COVID-19 had 3.19 (95% CI 2.63-3.88) times greater odds of in-hospital mortality compared to patients with AMI but no COVID-19. Patients with AMI and COVID-19 were also more likely to require mechanical ventilation (aOR 1.90, 95% CI 1.54-2.33), vasopressor use (aOR 1.60, 95% CI 1.11-2.33), and hemodialysis initiation (aOR 1.38, 95% CI 1.05-1.89) than patients with AMI without COVID-19. Patients with AMI and COVID-19 were also less likely to receive PCI (aOR 0.78, 95% CI 0.67 - 0.91), PCI or thrombolytics (aOR 0.80, 95% CI 0.69 – 0.93), and CABG surgery (aOR 0.40, 95% CI 0.28-0.59). However, they were more likely to receive thrombolytics (aOR 2.05, 95% CI 1.08- 3.92) compared to patients with AMI without COVID-19 (Table 3, Figure 2).

**Figure 2:**
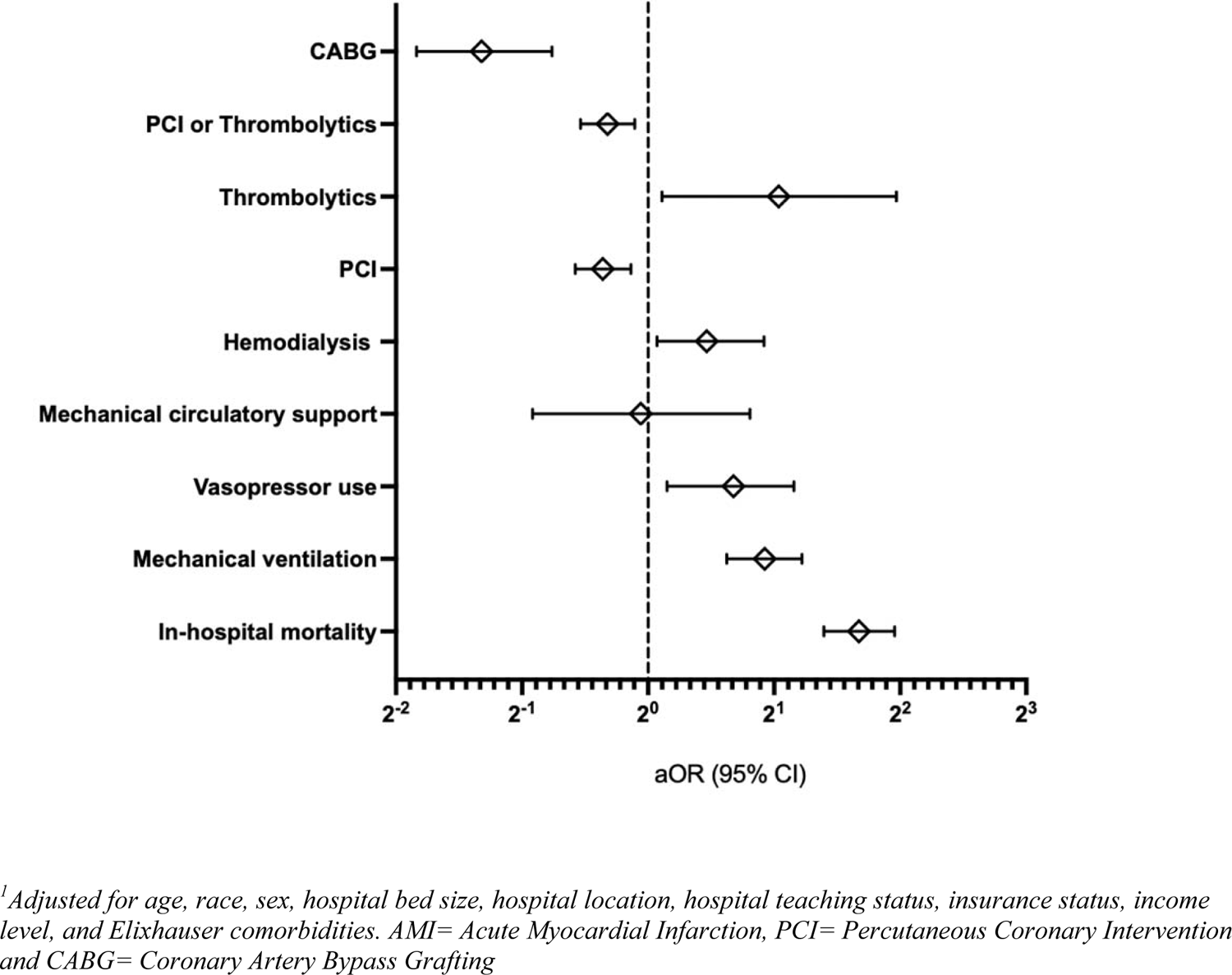
Outcomes, treatments and revasculazation odds for patients with COVID-19 and AMI relative to patients with AMI and no COVID-19 infection, using multivariate analysis

**Table 3:** Adjusted odds ratio outcomes for patients with COVID-19 and AMI relative to patients with AMI and no COVID-19 infection, using multivariate analysis.

|  | Adjusted odds ratio | P-value |
| --- | --- | --- |
| In-hospital mortality | 3.19 (95% CI 2.63 - 3.88) | < 0.01 |
| Mechanical ventilation | 1.90 (95% CI 1.54 - 2.33) | < 0.01 |
| Vasopressor use | 1.60 (95% CI 1.11 - 2.23) | < 0.01 |
| Mechanical circulatory support | 0.96 (95% CI 0.53 – 1.75) | 0.65 |
| Hemodialysis | 1.38 (95% CI 1.05 – 1.89) | 0.046 |
| PCI | 0.78 (95% CI 0.67 - 0.91) | < 0.01 |
| Thrombolytics | 2.05 (95% CI 1.08 – 3.92) | 0.028 |
| PCI or Thrombolytics | 0.80 (95% CI 0.69 – 0.93) | 0.003 |
| CABG | 0.40 (95% CI 0.28-0.59) | < 0.001 |

The in-hospital mortality rates for patients experiencing both AMI and COVID-19 showed variation across different racial groups. An analysis of the unadjusted clinical outcomes for racial differences was conducted. Compared to White patients with AMI and COVID-19; Black, Asian/Pacific Islander, and Native American patients experienced higher in-hospital mortality rates (p < 0.05) and increased hemodialysis (p < 0.001). Furthermore, Black, Hispanic, and Asian/Pacific Islander patients had lower rates of PCI (p<0.05) and lower rates of combined PCI or thrombolytics (p<0.05). (Supplemental Table 1, Supplemental Figure 2).

Upon adjusting for confounders, Black and Asian/Pacific Islander patients exhibited higher odds of in-hospital mortality in comparison to White patients, (aOR 2.13, 95% CI 1.35-3.59) and (aOR 3.41, 95% CI 1.5-8.37), respectively. Additionally, Black, Hispanic, and Asian/Pacific Islander patients showed higher odds of initiating hemodialysis, with odds ratios of (aOR 5.48, 95% CI 2.13-14.1), (aOR 2.99, 95% CI 1.13-7.97) and (aOR 7.84, 95% CI 1.55-39.5), respectively.

With regards to revascularization and after adjustment for cofounders, these patient groups still had significantly lower odds of receiving PCI compared to White patients: Black (aOR 0.71, 95% CI 0.67-0.74), Hispanic (aOR 0.81, 95% CI 0.77-0.86), and Asian/Pacific Islander (aOR 0.82, 95% CI 0.75-0.90). Moreover, Black patients had lower odds of undergoing CABG surgery compared to White patients (aOR 0.55, 95% CI 0.49-0.61) (Supplemental Table 2 and Supplemental Figure 3).

A subgroup analysis was performed to examine revascularization rates among patients diagnosed with STEMI. The baseline demographic characteristics for this subgroup can be found in (Supplemental Table 3). In patients with both STEMI and COVID-19, the unadjusted rate of undergoing PCI was lower (61.2% vs. 68.9%, p < 0.01), the rate of receiving thrombolytics was higher (2.6% vs. 0.9%, p < 0.01), and the rate of receiving either PCI or thrombolytics was lower (61.2% vs. 67.9%, p < 0.01) compared to patients with STEMI without COVID-19. After adjusting for confounding variables, patients with both STEMI and COVID-19 exhibited lower odds of undergoing PCI (aOR 0.73, 95% CI 0.58-0.91) and higher odds of receiving thrombolytic therapy (aOR 3.23, 95% CI 1.69-6.14). Meanwhile, the odds of receiving either PCI or thrombolytic therapy were lower (aOR 0.77, 95% CI 0.62-0.96) when compared to patients diagnosed with STEMI without COVID-19 (Table 4 and Figure 3).

**Figure 3:**
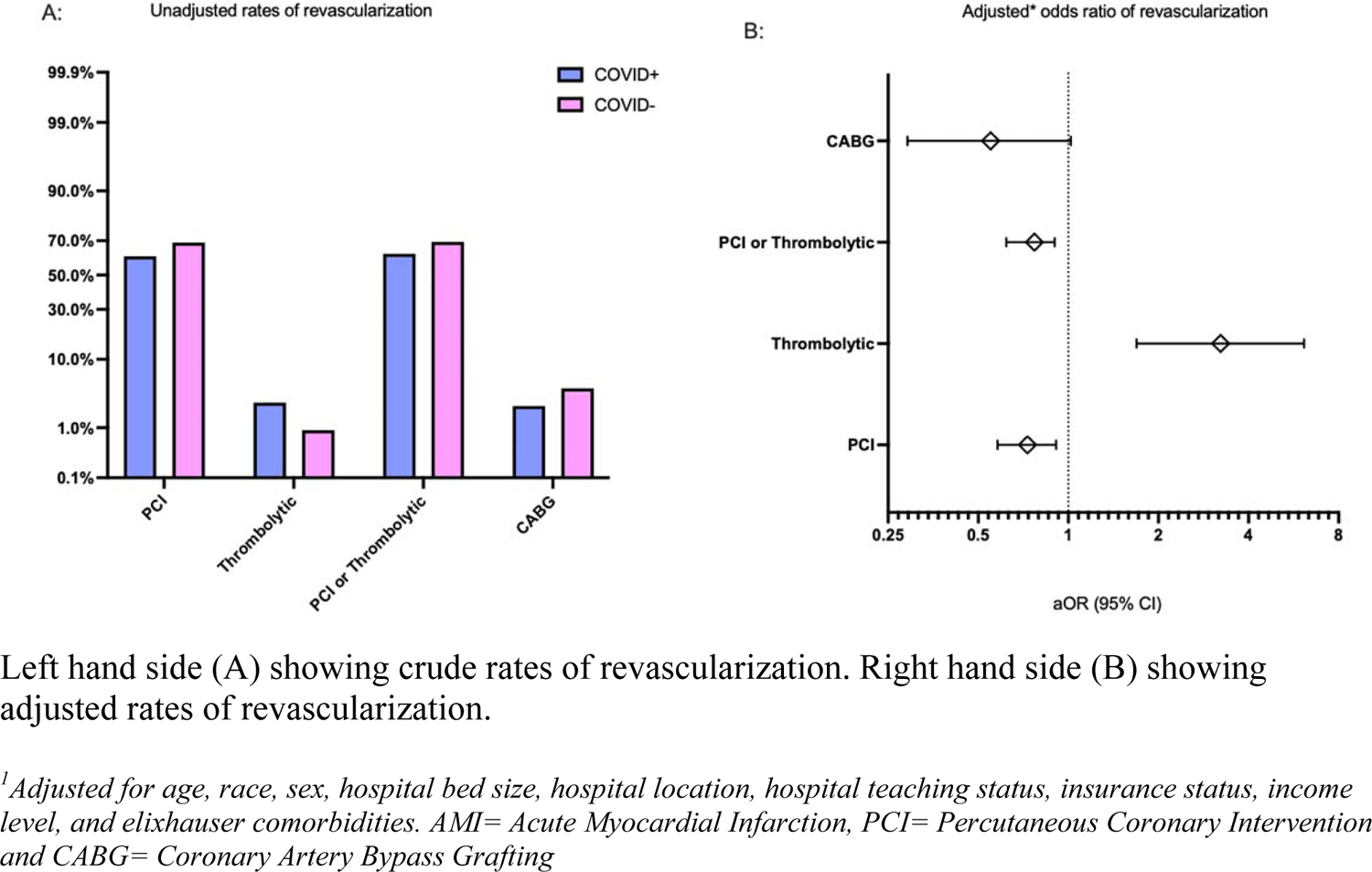
Rate of Revascularization with PCI, thrombolytics and CABG for STEMI + COVID-19 positive vs COVID-19 negative. Crude and Adjusted mortality rate both presented.

**Central Figure:**
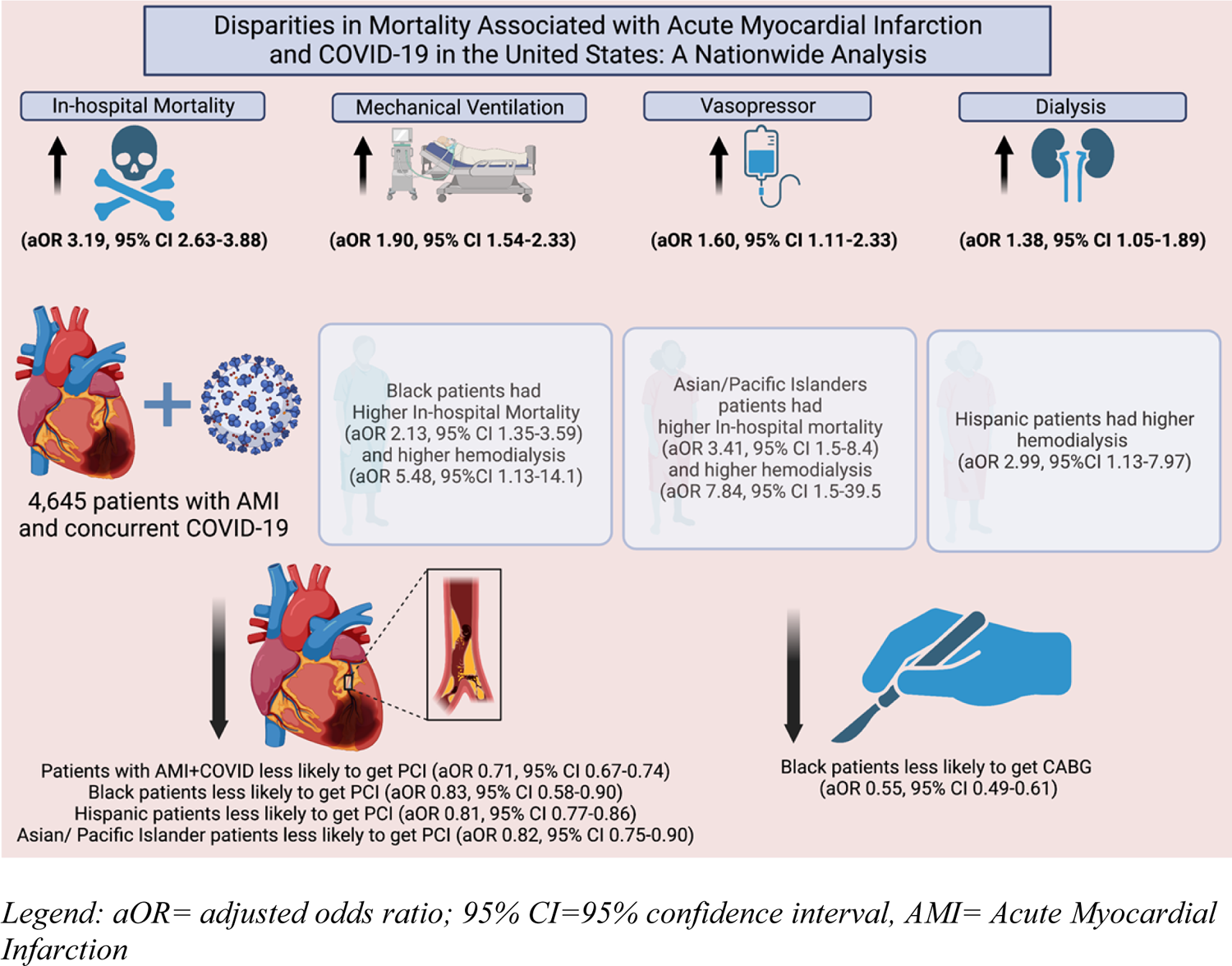
Disparities in Mortality Associated with Acute Myocardial Infarction and COVID-19 in the United States: A Nationwide Analysis

**Table 4:**
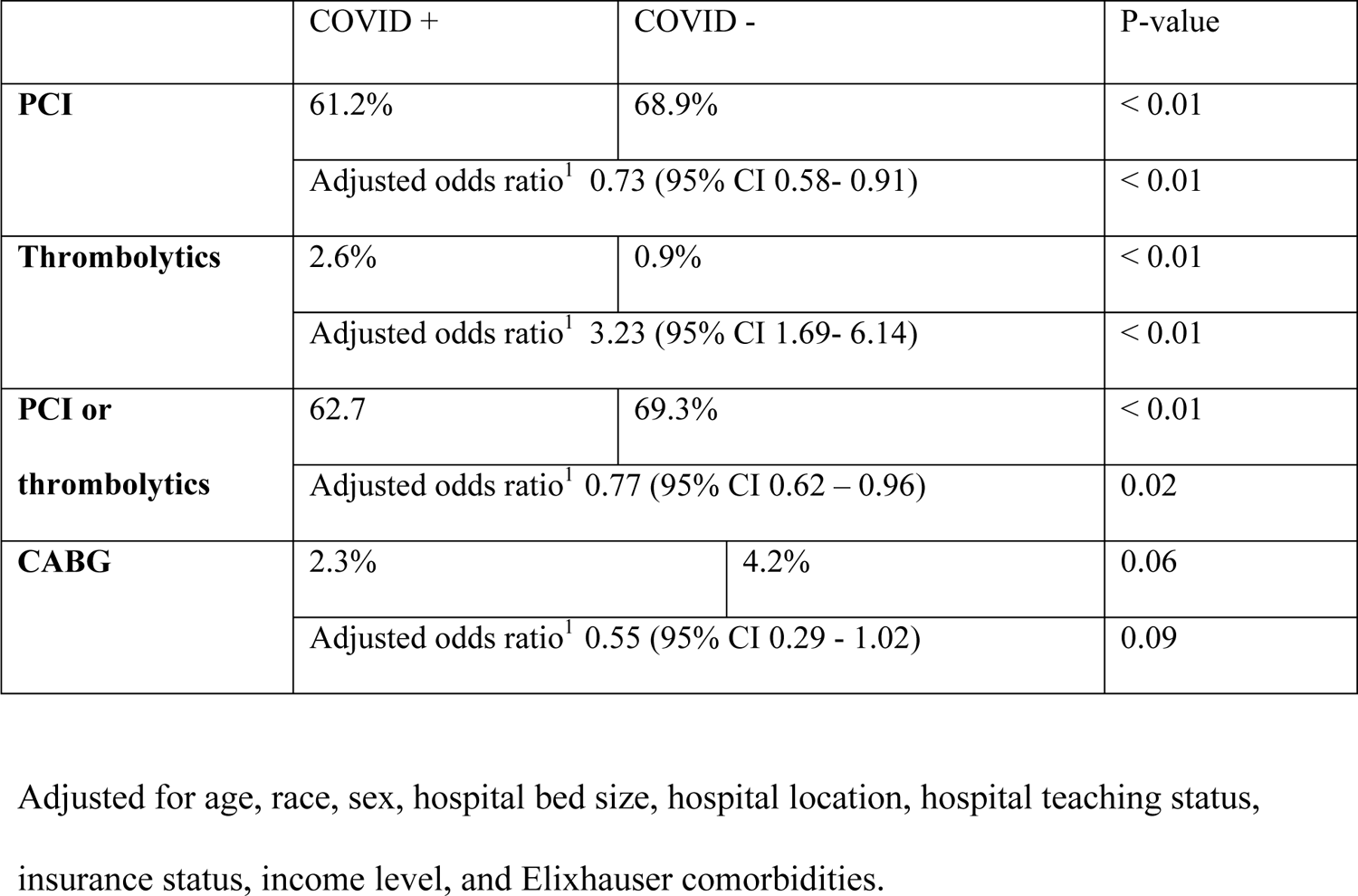
Rate of Revascularization with PCI, thrombolytics and CABG for STEMI + COVID-19 positive vs COVID-19 negative. Crude and Adjusted mortality rate both presented.

|  | COVID + | COVID - | P-value |
| --- | --- | --- | --- |
| **PCI** | 61.2% | 68.9% | < 0.01 |
|  | Adjusted odds ratio<sup>1</sup> 0.73 (95% CI 0.58- 0.91) |  | < 0.01 |
| **Thrombolytics** | 2.6% | 0.9% | < 0.01 |
|  | Adjusted odds ratio<sup>1</sup> 3.23 (95% CI 1.69- 6.14) |  | < 0.01 |
| **PCI or**<br>**thrombolytics** | 62.7 | 69.3% | < 0.01 |
|  | Adjusted odds ratio<sup>1</sup> 0.77 (95% CI 0.62 – 0.96) |  | 0.02 |
| **CABG** | 2.3% | 4.2% | 0.06 |
|  | Adjusted odds ratio<sup>1</sup> 0.55 (95% CI 0.29 - 1.02) |  | 0.09 |

Adjusted for age, race, sex, hospital bed size, hospital location, hospital teaching status, insurance status, income level, and Elixhauser comorbidities.

Rates of revascularization were furthermore analyzed by race. The unadjusted rates of revascularization revealed that Black, Hispanic, and Asian/ Pacific Islanders had lower rates of PCI (p<0.05) and lower rates of combined PCI or thrombolytics (p<0.05) (Supplemental Table 4). After accounting for confounding factors, Black, and Asian/Pacific Islander patients exhibited significantly lower odds of receiving PCI compared to White patients, (aOR 0.83, 95% CI 0.58- 0.90), and (aOR 0.78, 95% CI 0.66-0.90), respectively. Additionally, Black patients had lower odds of undergoing CABG surgery relative to White patients (aOR 0.68, 95% CI 0.53-0.87) (Supplemental Table 5).

## Discussion

This report examines the clinical management and outcomes of patients with AMI who were also diagnosed with COVID-19 in the United States during the first year of the pandemic. By examining AMI-related hospitalizations nationwide, it provides unique insights into the impact of COVID-19 on patients with AMI. The findings revealed a greater than three-fold increase in in-hospital mortality for patients with concurrent COVID-19 and AMI in comparison to those with only AMI. Several factors may have contributed to this increased mortality, including potential disruptions in care due to the presence of COVID-19 and the impact of the virus on the cardiovascular system. A recent large-scale observational study conducted with patients utilizing the National Health Service in England found that individuals diagnosed with both acute coronary syndrome (ACS) and COVID-19 were less likely to receive guideline-directed treatment and had a higher in-hospital and 30-day mortality rate compared to those without COVID-19 and ACS.^19^ Furthermore, data published from over 1300 chest pain centers in China showed an average delay of 20 minutes for reperfusion therapy early in the pandemic, which led to higher rates of in-hospital mortality and heart failure.^6^

Previous studies have highlighted a higher risk of complications and mortality for individuals with preexisting cardiovascular disease and COVID-19 infection.^20^ This analysis builds on this knowledge by demonstrating that patients with AMI and concurrent COVID-19 are more likely to require mechanical ventilation, vasopressor use, and initiation of hemodialysis. Notably, one study demonstrated that patients diagnosed with both ACS and COVID-19 infection have a higher incidence of pulmonary edema and shock at presentation, along with elevated troponin concentrations.^19^ Additionally, a Chinese study reported an 8% risk of acute cardiac injury, with a 13-fold higher incidence of cardiac injury in critically ill patients with COVID-19.^21^ Furthermore, patient concerns regarding COVID-19 exposure and avoidance of hospital visits may contribute to the increased mortality and complications associated with AMI during the pandemic. These factors raise concerns that untreated consequences of AMI may result in severe complications for many patients.^22–24^

The use of revascularization techniques, including PCI, thrombolytic therapy, and CABG surgery, was compared between patients diagnosed with STEMI with concurrent COVID-19 infection, and those with STEMI only. The analysis showed that patients with STEMI and COVID-19 were 27% less likely to receive PCI, 3.2 times more likely to receive thrombolytic therapy, and 23% less likely to receive either PCI or thrombolytic when compared with patients with STEMI and no COVID-19. This suggests that COVID-19 can negatively impact the delivery of crucial interventions for AMI patients, leading to higher in-hospital mortality rates (aOR 3.19, 95% CI 2.63 - 3.88) for those with AMI and COVID-19 compared to those without COVID-19. The findings emphasize the urgency of developing effective management strategies for patients with COVID-19 and cardiovascular disease. During the COVID-19 pandemic, multiple international guidelines tried to establish a consensus on the optimal treatment approach for patients presenting with AMI. The Chinese Cardiac Society recommended medical management for most patients presenting with NSTEMI and thrombolysis for those with STEMI early on in the pandemic.^25^ While American and Canadian guidelines had recommended the use of thrombolysis as an alternative to PCI for STEMI patients in cases where PCI services were limited.^6, 26^ Nonetheless, the ACC/SCAI and the European Association of Percutaneous Cardiovascular Interventions (EAPCI) recommendations encouraged the use of PCI as first-line therapy for STEMI.^26, 27^ Additionally a study done in Japan investigated the effect of the COVID- 19 pandemic on cardiovascular care, specifically analyzing hospital arrival time, ambulance use, PCI implementation, and in-hospital mortality. The results demonstrated no significant differences in these parameters before and after the outbreak. ^28^

Importantly the current analysis uncovered racial disparities in revascularization rates among the study cohort of patients with AMI and concurrent COVID-19, particularly with respect to PCI and CABG after AMI. Specifically, Black, Hispanic, and Asian/Pacific Islander patients were less likely to receive PCI for AMI than White patients, and Black patients were less likely to undergo CABG surgery for AMI. It is important to note that prior research has identified racial disparities in COVID-19 outcomes, with racial minority groups at an increased risk for morbidity and mortality due to COVID-19.^29–31^ The study shows disparities in treatment for AMI in the setting of COVID-19, suggesting a possible explanation for the overall poor outcomes seen in these diverse populations. The reasons for disparities in care after AMI are complex and multifactorial, including the role of racism, systemic bias, and other social determinants of health.^32^

## Limitations

The data used in this analysis was sourced from the NIS, which may have some inherent biases. The NIS does not include outpatient mortality, potentially leading to an underestimation of mortality in COVID-19 cases associated with AMI. Moreover, the NIS lacks specific data on lab values, vital signs, and imaging findings, so the conclusions drawn were based solely on discharge diagnoses. It is also impossible to determine, using the NIS data, whether AMI occurred after a COVID-19 infection or vice versa; only the presence of both diagnoses during a single admission is known. Ascertainment bias may be present, as patients were not routinely tested for COVID-19 at the beginning of the pandemic, which could explain the low rates of COVID-19 patients with AMI in this study. However, the mortality rates for patients with both AMI and COVID-19 in this study align with those found in previous studies.^19^ Finally, both AMI and COVID-19 cases in our study were identified using ICD-10 codes, which are subject to errors. Nonetheless, the large sample size in this study likely helps mitigate the impact of potential coding errors.

## Conclusion

Patients who were diagnosed with both COVID-19 and AMI had higher mortality rates and were more likely to experience complications during their hospital stay than those with only AMI. Furthermore, our findings identified racial disparities among patients with AMI and COVID-19, with Black and Asian/Pacific Islander patients receiving lower rates of revascularization compared to White patients with AMI and COVID-19 (Central Figure). This highlights the urgent need to address these systemic healthcare disparities to improve health equity in diverse patient populations. It is imperative that healthcare policies and interventions are implemented to ensure that all patients have access to high-quality care, regardless of their race or ethnicity. This will require a multifaceted approach, including increasing access to healthcare in underserved communities, promoting culturally sensitive care, and addressing the root causes of health disparities through social and economic policies. These findings should inform future research and policy initiatives aimed at reducing health disparities and improving health outcomes for all patients.

## ABBREVIATIONS & ACRONYMS

ACS: acute coronary syndrome

AHRQ: agency for healthcare research and quality

AMI: acute myocardial infarction

CABG: coronary artery bypass grafting

HCUP: healthcare cost and utilization project

NIS: National Inpatient Sample

NSTEMI: non-ST-elevation myocardial infarction

PCI: percutaneous coronary intervention (PCI)

STEMI: ST-elevation myocardial infarction

## Data Availability

All data referenced in this manuscript, obtained from the National Inpatient Sample, is publicly accessible and available for research purposes.

---

**Supplemental Figure 1:**
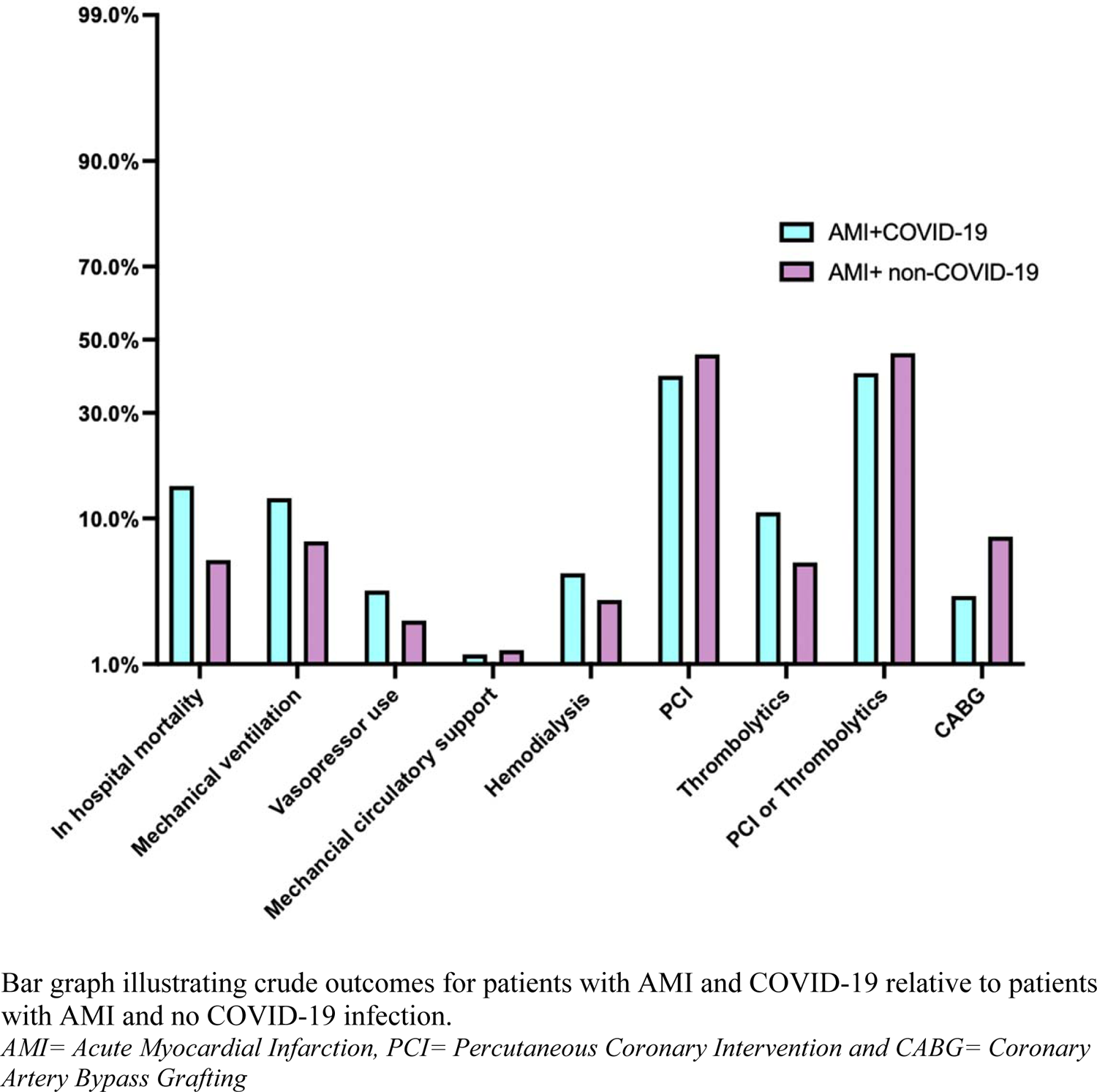
Outcomes, treatments and revascularization rates for patients with AMI and COVID-19 relative to patients with AMI and no COVID- 19.

**Supplemental Figure 2:**
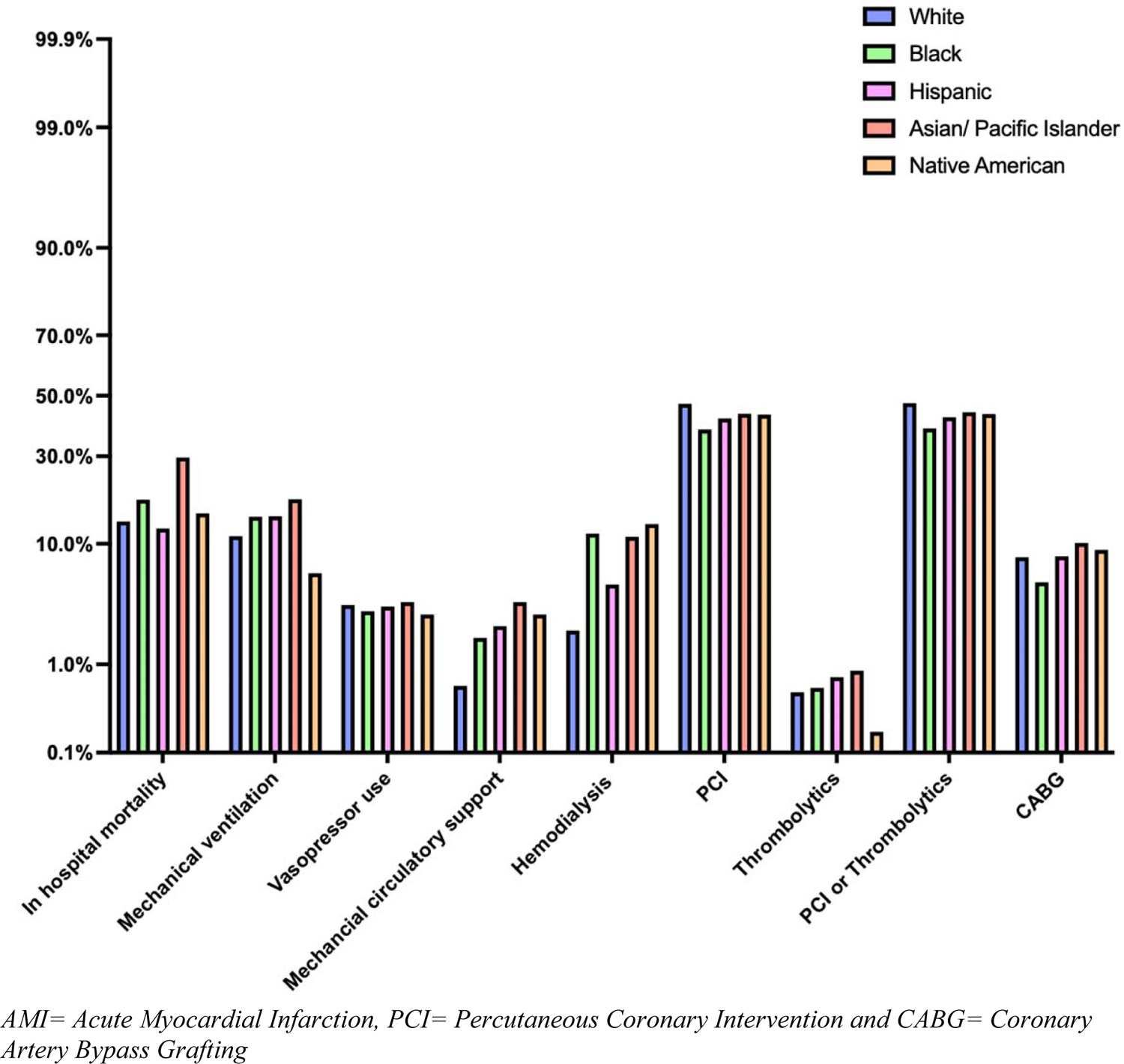
Unadjusted Outcomes, treatments, and revascularization rates of patients with AMI+ COVID-19 stratified by race.

**Supplemental Figure 3:**
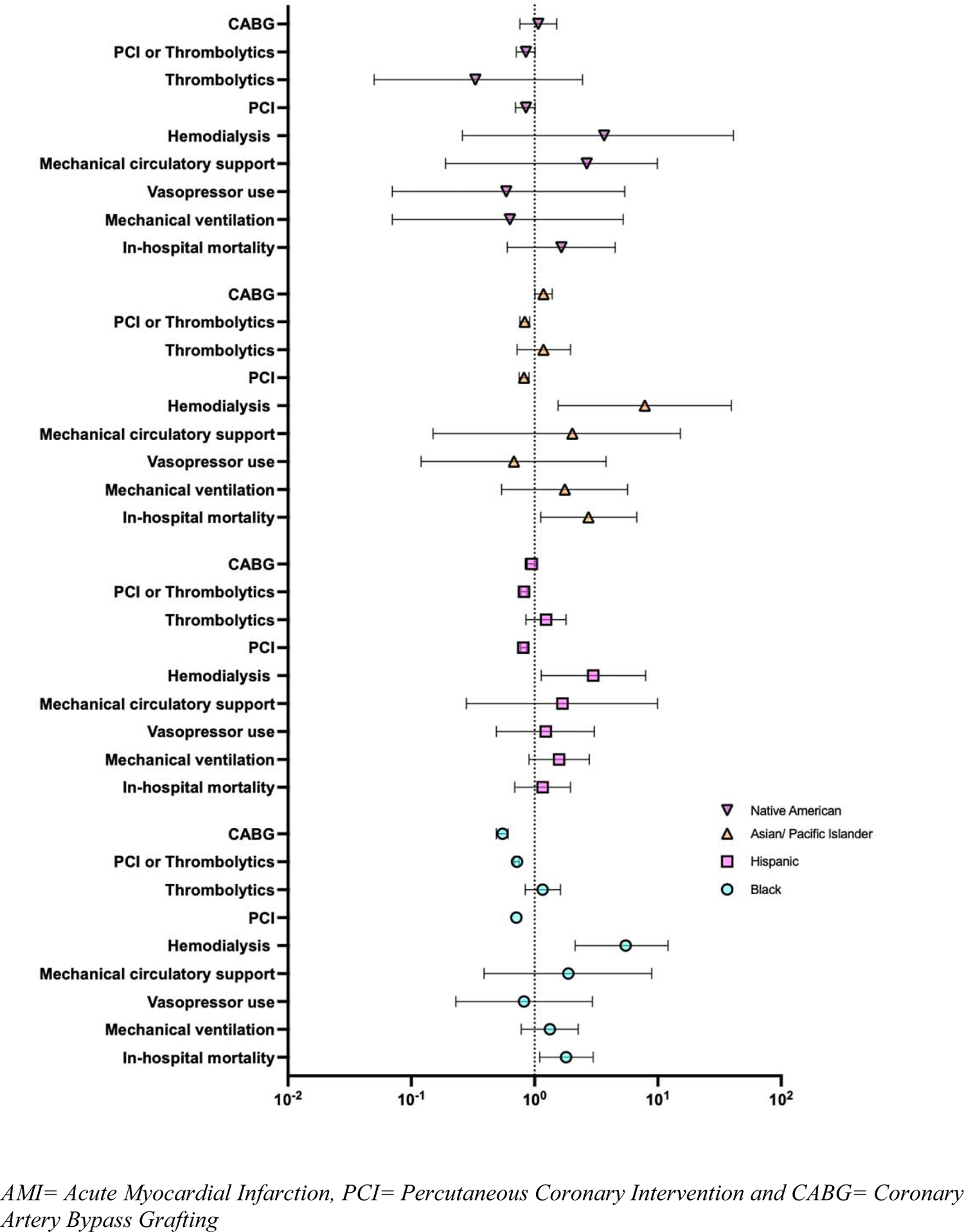
Forest plot illustrating outcomes for patients with COVID-19 and AMI relative to patients with AMI and no COVID-19 infection, stratified by race and using multivariate analysis.

**Supplemental Table 1:** Unadjusted primary and secondary outcomes of patients with AMI+ COVID-19 stratified by race.

|  |  |  |  | **Islander** | **American** |  |
| --- | --- | --- | --- | --- | --- | --- |
| In hospital mortality | 13.1% | 18.4% | 12.5% | 29.6% | 15.4% | 0.04 |
| Mechanical ventilation | 11.2% | 14.7% | 14.8% | 18.5% | 6.2% | 0.60 |
| Vasopressor use | 3.5% | 3.1% | 3.4% | 3.7% | 2.9% | 0.98 |
| Mechanical Circulatory Support | 0.6% | 1.8% | 2.3% | 3.7% | 2.9% | 0.37 |
| Hemodialysis | 2.1% | 11.6% | 5.1% | 11.1% | 6.3% | <0.001 |
| PCI | 47.2% | 38.5% | 42.2% | 43.8% | 43.5% | <0.001 |
| Thrombolytics | 0.51% | 0.57% | 0.74% | 0.86% | 0.18% | 0.067 |
| PCI or Thrombolytics | 47.4% | 38.8% | 42.6% | 44.3% | 43.7% | <0.001 |
| CABG | 8.1% | 5.3% | 8.2% | 10.1% | 9.1% | <0.001 |
| Mean length of stay (days) | 4.7 | 5.4 | 5.2 | 6.7 | 5.0 | 0.11 |
| Total Charges (USD) | 99,794 | 95,059 | 140,628 | 145,421 | 88,044 | 0.01 |

**Supplemental Table 2:**
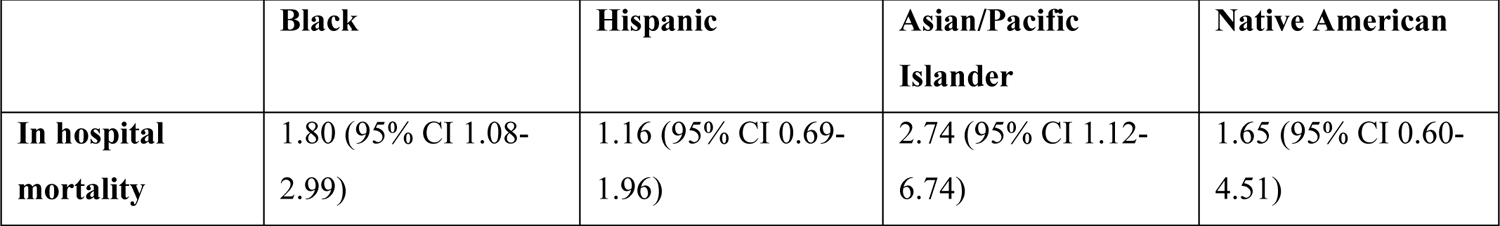
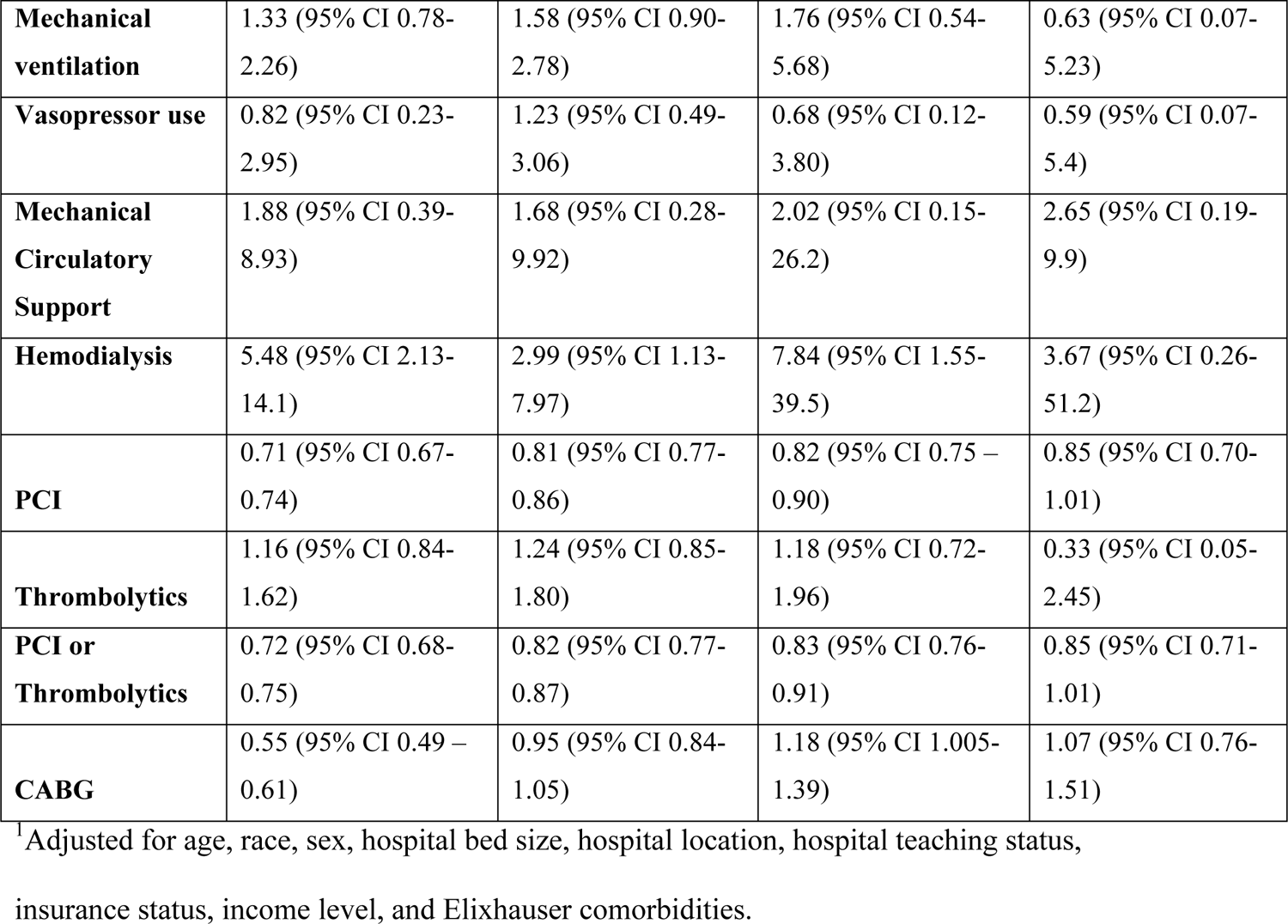
In-hospital mortality presented as adjusted odds ratio (aOR) for patients with AMI and COVID-19 distributed by race and relative to white race.

**Supplemental Table 3:** Baseline characteristics of all STEMI based on presence or absence of COVID-19.

| **Characteristics** | **STEMI+ COVID-19 (N=1945)** | **STEMI+ non-COVID-19 (N=139,325)** | **P-value** |
| --- | --- | --- | --- |
| (Female) | 30.6% (N=584) | 29.8% (N=48,012) | 0.72 |
| Mean Age | 61.8 | 62.9 | 0.12 |
| **RACE:** |  |  |  |
| White | 52.6% (N=1023) | 75.5% (N=112,226) | <0.001 |
| Black | 16.3% (N=332) | 8.9% (N=12,399) |  |
| Hispanic | 21.1% (N=317) | 8.4% (N=11,703) |  |
| Asian or Pacific Islander | 2.9% (N=56) | 3.0% (N=4,182) |  |
| Native American | 1.6% (N=31) | 0.5% (N=697) |  |
| **Median Household Income:** |  |  |  |
| \$ 1 - \$49,999 | 34.7% (N=674) | 27.9% (N=38,871) | 0.02 |
| \$ 50,000 - \$64,999 | 28.5% (N=554) | 28.6% (N=39,846) | |
| \$ 65,000 - 85,999 | 20.8% (N=405) | 23.5% (N=32,741) | |
| >\$86,000 | 16.0% (N=311) | 19.9% (N=27,745) | |
| **Insurance Status:** |  |  |  |
| Medicare | 36.5% (N=710) | 42.5% (N=59,213) | 0.01 |
| Medicaid | 15.7% (N=305) | 11.1% (N=15,465) |  |
| Private | 32.9% (N=640) | 34.8% (N=48,485) |  |
| Self-pay | 9.5% (N=185) | 7.3% (N=10,170) |  |
| No Charge | 1.03% (N=20) | 0.65% (N=891) |  |
| Other | 4.4% (N=86) | 3.6% (N=5016) |  |
| **Hospital Region:** |  |  |  |
| Northeast | 15.9% (N=309) | 16.6% (N=23,127) | 0.96 |
| Midwest | 23.7% (N=461) | 22.7% (N=31,626) |  |
| South | 40.1% (N=780) | 40.5% (N=56,426) |  |
| West | 20.3% (N=395) | 20.3% (N=28,282) |  |
| **Hospital Ownership:** |  |  |  |
| Government, nonfederal | 12.1% (N=235) | 9.1% (N=12,679) | 0.08 |
| Private, nonprofit | 75.3% (N=1465) | 75.5% (N=105,190) |  |
| Private, investor own | 12.6% (N=245) | 15.4% (N=21,456) |  |
| **Hospital Bedsize:** |  |  |  |
| Small | 22.1% (N=428) | 17.5% (N=24,382) | 0.03 |
| Medium | 24.7% (N=480) | 29.5% (N=41,100) |  |
| Large | 53.2% (N=1035) | 53.1% (N=73,981) |  |
| **Hospital Teaching Status:** |  |  |  |
| Rural non-teaching | 4.8% (N=93) | 6.1% (N=8,498) | 0.09 |
| Urban non-teaching | 15.4% (N=300) | 17.8% (N=24,800) |  |
| Urban Teaching | 79.7% (N=1550) | 76.1% (N=106,026) |  |
| **Elixhauser Comorbidity Index:** |  |  |  |
| 0 | 2.8% (N=55) | 4.3% (N=5,991) | 0.12 |
| 1 | 11.6% (N=226) | 14.1% (N=19,645) |  |
| 2 | 20.1% (N=391) | 20.8% (N=28,979) |  |
| >3 | 65.5% (N=1274) | 60.8% (N=84,710) |  |
| **Comorbidities:** |  |  |  |
| Atrial Fibrillation | 14.1% (N=274) | 13.1% (N=18,252) | 0.55 |
| Congested Heart Failure | 26.7% (N=519) | 26.3% (N=36,642) | 0.84 |
| Coronary Artery Disease | 81.2% (N=1579) | 86.2% (N=120,098) | 0.005 |
| Hypertension | 45.2% (N=879) | 46.3% (N=64,507) | 0.68 |
| Chronic Kidney Disease | 12.9% (N=251) | 10.8% (N=15,047) | 0.18 |
| Diabetes Mellitus | 41.7% (N=811) | 31.7% (N=44,166) | <0.001 |
| Hyperlipidemia | 59.9% (N=1165) | 68.1% (N=94,890) | <0.001 |
| Liver Disease | 1.3% (N=25) | 2.0% (N=2,786) | 0.32 |
| Lung Disease | 9.7% (N=187) | 10.9% (N=15,186) | 0.48 |
| Ischemic Stroke | 2.3% (N=45) | 0.9% (N=1,254) | 0.009 |
| Pulmonary HTN | 1.8% (N=35) | 2.5% (N=3,483) | 0.35 |
| Tobacco use | 35.9% (N=698) | 53.4% (N=74,400) | <0.001 |
| Obesity | 16.2% (N=315) | 16.2% (N=22,570) | 0.99 |

**Supplemental Table 4:** Unadjusted rates of revascularization for STEMI with concurrent COVID-19 with PCI, thrombolytics, and CABG stratified by race.

|  | White | Black | Hispanic | Asian/Pacific<br>Islander | Native<br>American | P-value |
| --- | --- | --- | --- | --- | --- | --- |
| **PCI** | 69.7% | 65.2% | 66.7% | 64.6% | 71.2% | < 0.05 |
| **Thrombolytics** | 0.9% | 1.0% | 1.0% | 1.1% | 0.7% | 0.98 |
| **PCI or** | 70.0% | 65.8% | 67.1% | 64.9% | 72.3% | < 0.05 |
| **thrombolytics** |  |  |  |  |  |  |
| **CABG** | 4.2% | 3.2% | 4.9% | 4.7% | 5.1% | 0.06 |

**Supplemental Table 5:** Adjusted^1^ rates of revascularization with PCI, thrombolytics and CABG stratified by race and relative to White race, presented as odds ratios.

|  | **Black** | **Hispanic** | **Asian/Pacific**<br>**Islander** | **Native American** |
| --- | --- | --- | --- | --- |
| **PCI** | 0.83 (95% CI 0.58-0.90) | 0.91 (95% CI 0.83-1.00) | 0.78 (95% CI 0.66-0.90) | 1.05 (95% CI 0.71-1.5) |
| **Thrombolytics** | 1.09 (95% CI 0.65-1.85) | 0.94 (95% CI 0.57-1.55) | 0.94 (95% CI 0.46-1.90) | 0.77 (95% CI 0.10-5.70) |
| **PCI or** | 0.83 (95% CI 0.75- | 0.91 (95% CI 0.82- | 0.77 (95% CI 0.66- | 1.07 (95% CI 0.72- |
| **thrombolytics** | 0.92) | 1.00) | 0.90) | 1.58) |
| **CABG** | 0.68 (95% CI 0.53-0.87) | 1.13 (95% CI 0.91-1.40) | 1.11 (95% CI 0.79-1.58) | 1.10 (95% CI 0.49-2.45) |

<sup>1</sup>Adjusted for age, race, sex, hospital bed size, hospital location, hospital teaching status, insurance status, income level, and Elixhauser comorbidities.

